# Mapping associations of polygenic scores with autism and ADHD traits in a single city region

**DOI:** 10.1101/2022.09.22.22280240

**Authors:** Zoe E. Reed, Richard Thomas, Andy Boyd, Gareth J. Griffith, Tim T. Morris, Dheeraj Rai, David Manley, George Davey Smith, Oliver S.P. Davis

## Abstract

**Background:** The genetic and environmental aetiology of autistic and Attention Deficit Hyperactivity Disorder (ADHD) traits is known to vary spatially, but does this translate into variation in the association of specific common genetic variants?

**Methods:** We mapped associations between polygenic scores for autism and ADHD and their respective traits in the Avon Longitudinal Study of Parents and Children (N=4,255 to 6,165) across the area surrounding Bristol, UK, and compared them to maps of environments associated with the prevalence of autism and ADHD.

**Results:** Our maps suggest genetic associations vary spatially, with consistent patterns for autistic traits across polygenic scores constructed at different p-value thresholds. Patterns for ADHD traits were more variable across thresholds. We found that the spatial distributions often correlated with known environmental influences.

**Conclusions:** These findings shed light on the factors that contribute to the complex interplay between the environment and genetic influences in autism and ADHD traits.

**Key points:**

- The prevalence of autism and ADHD vary spatially.
- Our study highlights that genetic influences based on PGS also vary spatially.
- This spatial variation correlates with spatial variation in environmental characteristics as well, which would be interesting to examine further.
- Our findings have implications for future research in this area examining the factors that contribute to the complex interplay between the environment and genetic influences on autistic and ADHD traits.

## Introduction

The prevalence of both autism and Attention Deficit Hyperactivity Disorder (ADHD) is known to vary by location (Arns, Van Der Heijden, Arnold, & Kenemans, 2013; Chiarotti & Venerosi, 2020; Delobel-Ayoub et al., 2020; Hoffman et al., 2017; Vieira, Fabian, Webster, Levy, & Korrick, 2017). For example, both more commonly occur in areas of greater urbanicity, although evidence for this is less clear for ADHD than for autism (Chen, Liu, Su, Huang, & Lin, 2008; Marlene B. Lauritsen et al., 2014; Madsen, Ersbøll, Olsen, Parner, & Obel, 2015; Markevych et al., 2014a; Wu & Jackson, 2017). Autism appears to be more prevalent in areas with greater average socioeconomic position (SEP) and more readily available diagnostic services (Bakian, Bilder, Coon, & McMahon, 2015; Mazumdar, Winter, Liu, & Bearman, 2013; Van Meter et al., 2010). Some studies suggest lower ADHD prevalence in areas with greater solar intensity (Arns et al., 2013; Arns, Swanson, & Arnold, 2018), in line with evidence that lower vitamin D levels are associated with increased ADHD risk (Khoshbakht, Bidaki, & Salehi-Abargouei, 2018). Both traits also show strong genetic influence, with heritability estimated at around 80% (Faraone & Larsson, 2018; Larsson, Chang, D’Onofrio, & Lichtenstein, 2014; Rietveld, Hudziak, Bartels, van Beijsterveldt, & Boomsma, 2004; Tick, Bolton, Happé, Rutter, & Rijsdijk, 2016). Recent genome-wide association studies (GWAS) of autism (Grove et al., 2019) and ADHD (Demontis et al., 2019) have confirmed both are highly polygenic. Polygenic scores (PGS) for autism and ADHD constructed from associated variants have been shown to predict autistic and ADHD traits in other populations (Burton et al., 2018; M. J. Taylor et al., 2019).

It is currently unclear whether this genetic aetiology varies spatially in a similar way to prevalence. Previous research on autistic traits using twin data suggests there is broad spatial variation within countries in genetic and environmental influences (Davis, Haworth, Lewis, & Plomin, 2012; Reed et al., 2021). However, we do not yet know whether similar variation is apparent at higher spatial resolution within a single city region, or using known genetic variants associated with autism and ADHD.

Here we used variants from the GWAS described above to construct PGS for participants in the Avon Longitudinal Study of Parents and Children (ALSPAC), a geographically clustered birth cohort. We conducted weighted analyses across a regular grid of spatial points covering the area surrounding the city of Bristol in the United Kingdom (UK), to examine high resolution spatial variation in associations between PGS and autism and ADHD traits.

## Methods

### Cohort description

ALSPAC initially recruited 14,541 pregnant women resident in the former county of Avon centred on the city of Bristol, UK with expected delivery dates between 1st April 1991 and 31st December 1992. Of these initial pregnancies, 13,988 children were alive at age 1. When the children were approximately age 7, additional eligible cases who had failed to join the study originally were recruited, resulting in a total sample size of 14,901 children (Boyd et al., 2013, 2019; Fraser et al., 2013). The study website contains details of all the data that is available through a fully searchable data dictionary and variable search tool (http://www.bristol.ac.uk/alspac/researchers/our-data/).

Ethical approval for the study was obtained from the ALSPAC Ethics and Law Committee and the Local Research Ethics Committees. Informed consent for the use of data collected via questionnaires and clinics was obtained from participants following the recommendations of the ALSPAC Ethics and Law Committee at the time. Consent for biological samples has been collected in accordance with the Human Tissue Act (2004). Participants included in our analyses were restricted to those residing in the area in and around Bristol when measures were obtained.

### Phenotypic measures

#### Attention-deficit hyperactivity disorder traits

We used parent responses on the Strengths and Difficulties Questionnaire hyperactivity/inattention subscale (Goodman, 1997), completed when children were a mean age of 9.64 (SD=0.12). This scale has good internal consistency (Cronbach’s alpha of 0.78) (Mieloo et al., 2012), test-retest reliability (0.81) (Stone et al., 2015), sensitivity (75%) (Goodman, Ford, Richards, Gatward, & Meltzer, 2000) and specificity (84%) (Hall et al., 2019) for ADHD diagnosis. It can sufficiently distinguish between clinical and community samples (Vugteveen, de Bildt, Theunissen, Reijneveld, & Timmerman, 2021). The scale consists of the following five items: ‘Restless, overactive, cannot stay still for long’, ‘Constantly fidgeting or squirming’, ‘Easily distracted, concentration wanders’, ‘Think things out before acting’ (reverse scored), ‘Sees tasks through to the end. Good attention span’ (reverse scored). Responses are scored as ‘Not true’ (0), ‘Somewhat true’ (1) and ‘Certainly true’ (2), with a maximum total score of 10. The score distribution is presented in Supplementary Fig S1.

#### Autistic traits

We used two measures of autistic traits. The first, which we refer to as social autistic traits, was administered on a single occasion, at a mean age of 10.72 (SD=0.12). We used total scores from parent responses to the Social and Communication Disorders Checklist (SCDC) (Skuse, Mandy, & Scourfield, 2005). The SCDC has high internal consistency (Cronbach’s alpha of 0.93), test-retest reliability (0.81), sensitivity (90%) and specificity (69%) for an autism diagnosis (Skuse et al., 2005). It consists of 12 items, with responses scored as ‘Not true’ (0), ‘Quite/Sometimes true’ (1), and ‘Very/Often true’ (2), with a maximum total score of 24. The score distribution is presented in Supplementary Fig S2.

The second measure, which we refer to as the autistic traits mean factor score, was derived from 93 measures (including SCDC measurements) obtained at multiple time points from age 6 months to 9 years (Steer, Golding, & Bolton, 2010). The SCDC measure is a more specific measure of autistic social traits, but the autistic traits mean factor score encompasses a broader measure of autistic traits, and provides a useful test of the sensitivity of the results to changes in phenotypic measurement. Further details can be found in the Supporting Information (Section 1 and Fig S3). We flipped the sign of the score so that a more positive score corresponds to a stronger indication of autistic traits. The phenotypic correlation between the two measures was 0.44.

#### Covariates

We included the child’s sex and age at assessment as covariates in analyses of social autistic and ADHD traits. For the autistic traits mean factor score we included sex, but not age since the score is a composite of measures at multiple time points. We also included the first 20 principal components (PCs) of population structure in our unweighted analyses to assess whether this may influence our findings.

#### Location data and weightings

We conducted analyses at a regular hexagonal grid of 1036 locations (see Supplementary Fig S4) across the ALSPAC recruitment area, comprising the three health districts that existed in the old county of Avon (Southmead, Frenchay, and Bristol and Weston District Health Authorities). This spatial resolution was chosen as it allowed a good trade-off between greater resolution and the number of data points manageable for analysis in a multi-step model where the ALSPAC team and the researchers exchanged datasets several times to allow the use of accurate spatial information from participants without it being released to researchers. See Supporting Information (Section 2) for further details.

Participants’ contributions to each analysis were weighted by a function of their Euclidean distance from the analysis location. Participants were assigned locations corresponding to the centroid of their residential postcode area at age 10. A postcode area groups a mean of 15 neighbouring properties and covers a mean area of 43,830m^2^. The weighting function is given below, where *x_i_* is the participant’s location, *x* is the analysis location, *d* is the Euclidean distance between these, and *w_i_* is the weight for each participant:

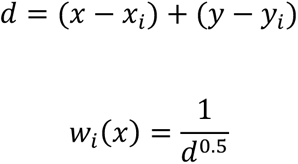

The power parameter we have used is 0.5 to allow for a trade-off between more accurate estimation of the association and accurately localising this, where estimates are smoothed somewhat towards population means whilst allowing for patterns of variation to be observed. This allowed each participant to contribute to each analysis, with participants living closer to an analysis location contributing greater weight to the analysis.

### Genetic data

Genetic data for children and mothers were obtained from a combination of blood and buccal samples (see Supporting Information Section 3). After quality control and removing those who had withdrawn consent, there were 8,252 children and 7,914 mothers with genotype data available.

### Polygenic score construction

The construction of PGS is described in detail in Supporting Information (Section 4). Briefly, we used Plink (version 2) (Purcell et al., 2007) to construct weighted PGS for each participant from GWAS summary statistics for ADHD (Demontis et al., 2019) and autism (Grove et al., 2019) by summing the number of risk alleles present for each SNP (0, 1 or 2) weighted by the effect of that SNP in the GWAS discovery sample. We generated maps for multiple PGS constructed at the *p*-value thresholds (*pT*) *p* <5×10^-8^, *p* <1×10^-5^, and *p* <0.5 in the discovery GWAS, and for the threshold that explained the most variance in the phenotype in the full, unweighted ALSPAC sample. We standardised PGS to *z* scores, so results are presented on the scale of standard deviation (SD) changes in PGS.

### Statistical analysis

All analyses were conducted in R (V3.6.2).

#### Spatial variation using weighted polygenic score analyses

Initially we conducted analyses without weighting by location to obtain estimates for the association of the PGS with the phenotypes (see Supporting Information Section 5). We then ran linear regression models for each of 1,036 locations, with participants’ contributions to each analysis weighted by the Euclidean distance from the analysis location. We compared the spatial distribution of results for different *pT* with the Lee statistic (spdep R package, version 1.1-2) (Bivand & Wong, 2018; S. Il Lee, 2001, 2004). This is a global bivariate spatial correlation test, which integrates an aspatial bivariate measure (Pearson’s correlation) and a univariate spatial measure (Moran’s I). It captures spatial co-patterning and therefore the extent to which bivariate associations are spatially clustered. Results are interpreted as the spatial similarity of the two distributions (a combination of the correlation between the measures and spatial clustering). We have no strong hypothesis about the direction of effect and results in either direction were of interest. Therefore, p-values reflect two-tailed tests.

#### Maps of environmental characteristics

We examined several environmental variables previously found to be associated with the prevalence of autism and ADHD, as described in the introduction: population density (Donovan, Michael, Gatziolis, Mannetje, & Douwes, 2019; Marlene B. Lauritsen et al., 2014; Marlene Briciet Lauritsen, Pedersen, & Mortensen, 2005; Madsen et al., 2015; Markevych et al., 2014a; Vassos, Agerbo, Mors, & Bøcker Pedersen, 2016; Wu & Jackson, 2017); parental education level, neighbourhood educational attainment and SEP (Bakian et al., 2015; Hoffman, Kalkbrenner, Vieira, & Daniels, 2012; Russell, Ford, & Russell, 2015; Vieira et al., 2017); and low exposure to sunlight (Arns et al., 2013; Hastie et al., 2019; Vinkhuyzen et al., 2018).

To assess whether these environmental characteristics were also correlated with differences in the strength of the association between polygenic scores for autism and ADHD and the phenotypes themselves, we created maps of each environmental measure over the same area, using data from external sources. We quantify these in our model by including measures of population density, average qualification level, level of urbanicity, the index of multiple deprivation (IMD) and hours of bright sunshine (see Supporting Information Section 6 and Supplementary Table 9). Data for population density and IMD were log transformed due to positive skews. We used the Lee statistic to compare the spatial distributions of these environmental variables to the maps of variation in PGS association.

#### Associations between polygenic scores and participation and migration measures

To index sampling bias, we tested the association between children’s and mothers’ PGS, participation rates and migration out of the Avon area. Loss to follow-up could be associated with PGS for autism and ADHD, as suggested previously (A. E. Taylor et al., 2018). To assess this, we created measures of each child’s and mother’s participation in ALSPAC, up to child age 11 (see Supporting Information Section 7). For analyses using mother’s PGS we adjusted for mother’s age.

### Data availability

ALSPAC data access is through a system of managed open access. Access can be applied for as detailed in the [ALSPAC access policy](https://www.bristol.ac.uk/media-library/sites/alspac/documents/researchers/data-access/ALSPAC_Access_Policy.pdf).

### Code availability

The analysis code used in this study is available upon request from the authors.

## Results

### Sample description

After excluding those without the phenotypic, location and genetic data (for PGS) required, we included between 4,255 and 6,165 children in each analysis (see Supplementary Table S1).

### Population-level polygenic score analysis

Results for population-level PGS analyses, with and without additional adjustment for 20 PCs, are presented in Supplementary Tables S2, S3 and S4 for ADHD traits, social autistic traits and the autistic traits mean factor score, respectively. When adjusting additionally for the 20 PCs, effects are attenuated. The *p-value thresholds* that explain the most variance are 0.5 for ADHD traits (*N*=5,258; *r^2^*=0.011), 0.1 for social autistic traits (*N*=5,200; *r^2^*=0.00068) and 0.5 for the autistic traits mean factor score (*N*=7,505; *r^2^*=0.0012). We have generated maps for these *pT* along with the other selected *pT*s, resulting in 3 analyses for ADHD traits and the autistic mean factor score and 4 analyses for social autistic traits (Figs 1-3).

**Fig 1.**
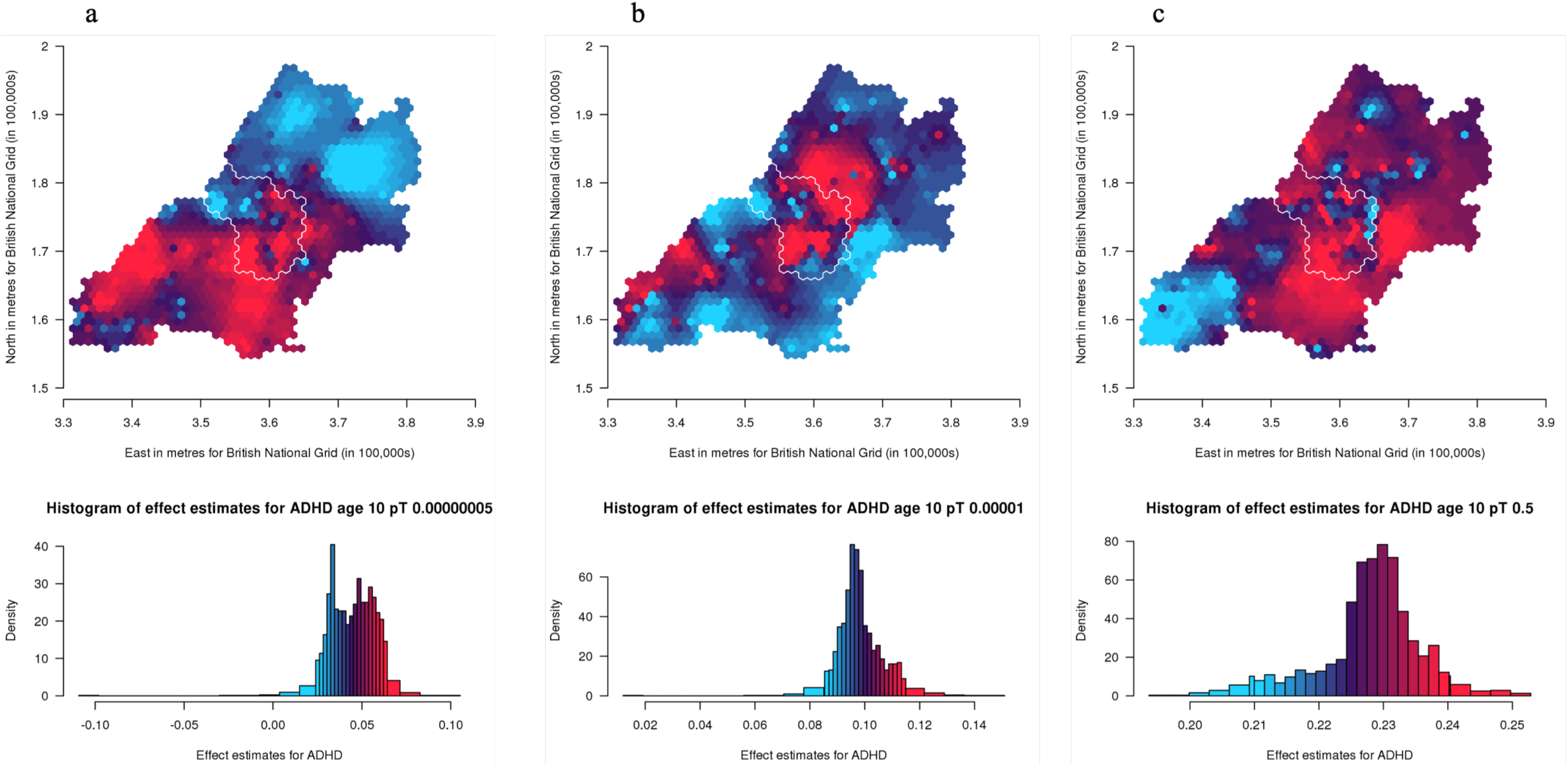
Mapping the association of the polygenic score for ADHD with ADHD traits shows a lack of consistency in results across different p-value thresholds over the area surrounding Bristol, UK *Spatial variation in genetic influences ranging from low (blue) to high (red). Histograms show the distribution of effect estimates, coloured in the same way. The city of Bristol is outlined in white*.

**Fig 3.**
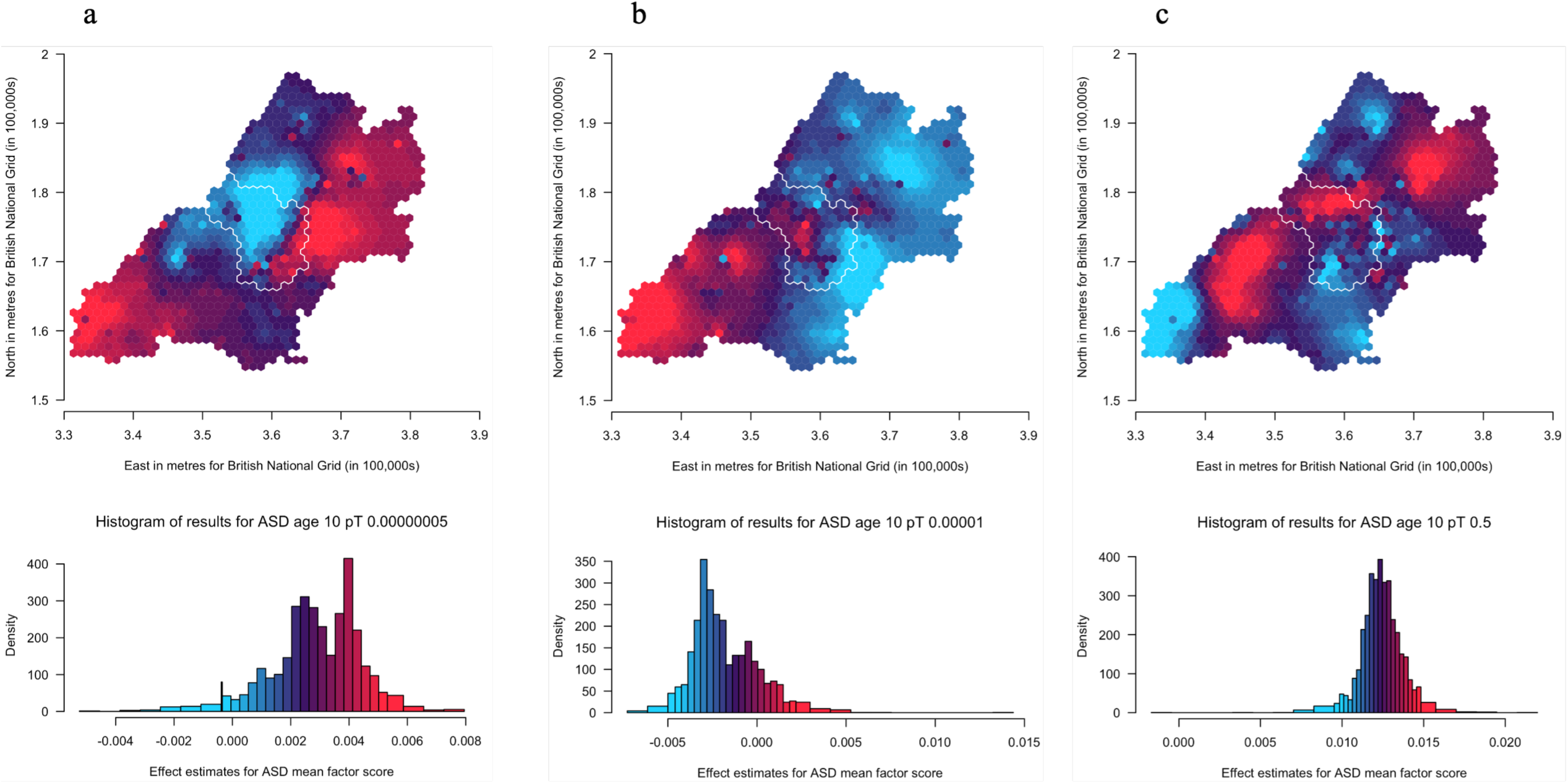
Mapping the association of the polygenic score for autism with the autistic traits mean factor score shows some consistency in variation across the p-value thresholds over the area surrounding Bristol, UK *Spatial variation in genetic influences ranging from low (blue) to high (red). Histograms show the distribution of effect estimates, coloured in the same way. The city of Bristol is outlined in white*.

### Spatially weighted polygenic score analyses

Maps of spatially weighted PGS for ADHD traits (N=4,309) are presented in Fig 1 (a-c) (*pT*: 5×10^-8^, 1×10^-5^ and 0.5, respectively). From visual inspection it is difficult to recognise patterns across the thresholds used. However, there are a few areas that appear more consistent, for example within Bristol, the north-west generally has lower genetic influence whilst the south has higher genetic influence.

Results for spatially weighted PGS for social autistic traits (N=4,255) are presented in the maps in Fig 2 (a-d) (pT: 5×10^-8^, 1×10^-5^, 0.1 and 0.5, respectively). These results appear more consistent across the different *pT* than for ADHD traits, even though the autistic traits PGS explains less variance than the ADHD PGS. We generally see higher effect estimates in the south-west and north-west of the region than in the east. Low estimates are also seen around the most south-west area and this is most apparent for the 0.5 *pT*. The area within the city of Bristol shows variation, with north-western areas of the city generally showing higher estimates compared to the south-eastern areas.

**Fig 2.**
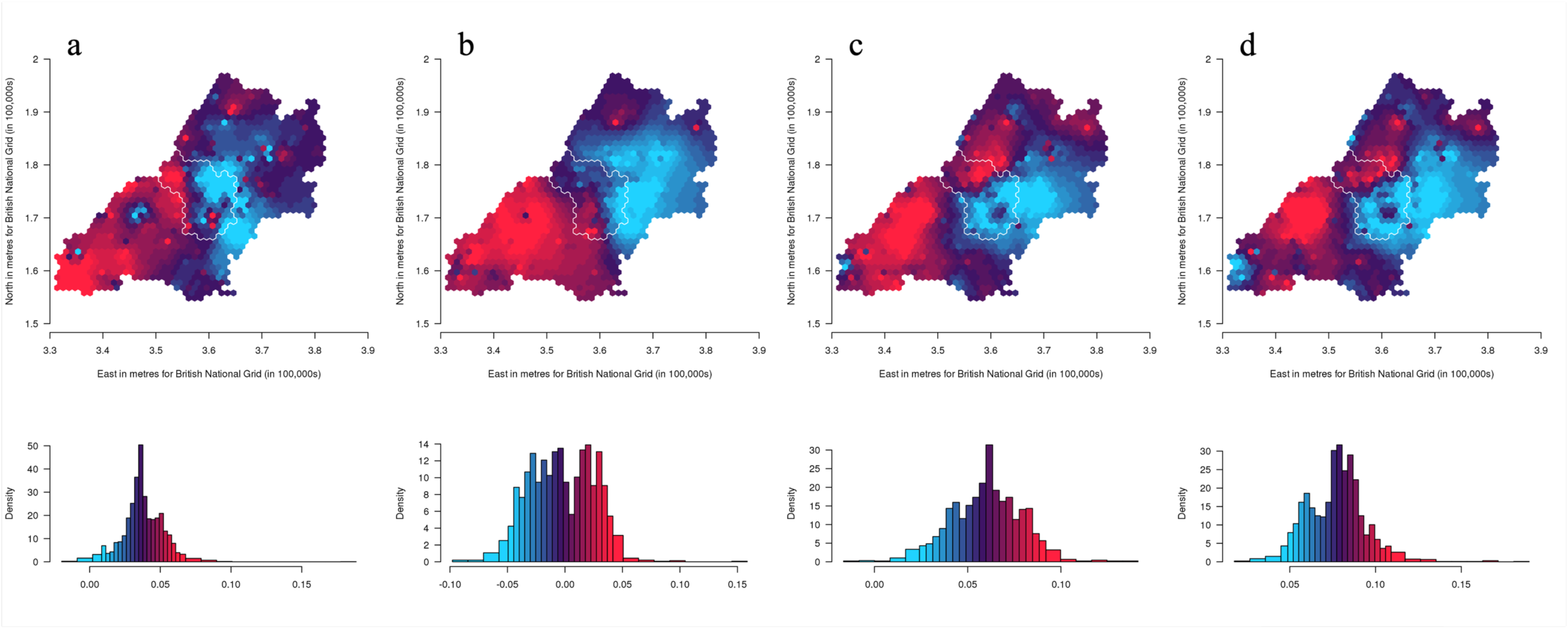
Mapping the association of the polygenic score for autism with social autistic traits shows consistent variation across the p-value thresholds over the area surrounding Bristol, UK *Spatial variation in genetic influences ranging from low (blue) to high (red). Histograms show the distribution of effect estimates, coloured in the same way. The city of Bristol is outlined in white*.

Results for spatially weighted PGS for the autistic traits mean factor score (N=6,165) are presented in the maps in Fig 3 (a-c) (pT: 5×10^-8^, 1×10^-5^ and 0.5, respectively). These results appear less consistent across the different *pT* than those for social autistic traits. However, there are some consistencies: the most south-westerly area, with a similar pattern to social autistic traits, is relatively higher at lower *pT* compared to other areas, and lower at the higher *pT*. The east has generally low values compared to the west and northern areas, similarly to social autistic traits. We also see within-city variation for Bristol, with the north-western areas showing higher estimates compared to the south-eastern areas of the city of Bristol at higher *pT*.

We compared maps across the different *pT* for each trait using Lee’s L statistic (Supplementary Table S5). As is apparent from visual inspection, results for ADHD traits are not strongly spatially correlated across *pT*, although maps for *pT* 5×10^-8^ and 1×10^-5^ are more correlated (Lee’s statistic=0.14, *p*=0.002) than maps for *pT* 1×10^-5^ and 0.5 (Lee’s statistic=-0.008, *p*=0.07). For social autistic traits we observe stronger associations across all *pT* (Lee’s statistic=0.57 to 0.81, *p*<2×10^-04^), confirming the observed spatial consistency in the patterns. For the autistic traits mean factor score, correlations are much weaker (Lee’s statistic=-0.22 to 0.07, *p*<2×10^-04^).

### Risk factor maps and comparison of spatial distributions

Maps of population density, average qualification level, IMD, level of urbanicity and hours of sunshine are shown in Fig 4 (a-e), respectively. Results for the Lee test comparing these maps with the PGS maps, at the *pT* explaining the most variance, are shown in Supplementary Table S6. To account for multiple testing, we applied a Bonferroni correction and considered a p-value <0.003 to be strong evidence of correlation. For ADHD traits, there is strong evidence of correlations with all environmental measures. Strong evidence of a positive correlation was found with average qualification level (Lee statistic=0.07, *p*<2×10^-04^) and negative correlations with the other measures (Lee statistic=-0.04 to -0.47, *p*<2×10^-04^), with the strongest correlation being with hours of sunshine. For social autistic traits, we found strong evidence of positive correlations with average qualification level (Lee statistic=0.07, p<2×10^-04^) and hours of sunshine (Lee statistic=0.57, p<2×10^-04^) and negative correlations with population density (Lee statistic=-0.11, *p*<2×10^-04^) and IMD (Lee statistic=-0.18, *p*<2×10^-04^). The autistic traits mean factor score showed strong evidence of a positive correlation with average qualification level (Lee statistic=0.13, p<2×10^-04^) and negative correlation with IMD, urbanicity and hours of sunshine (Lee statistic=-0.05 to -0.19, *p* <2×10^-04^).

**Fig 4.**
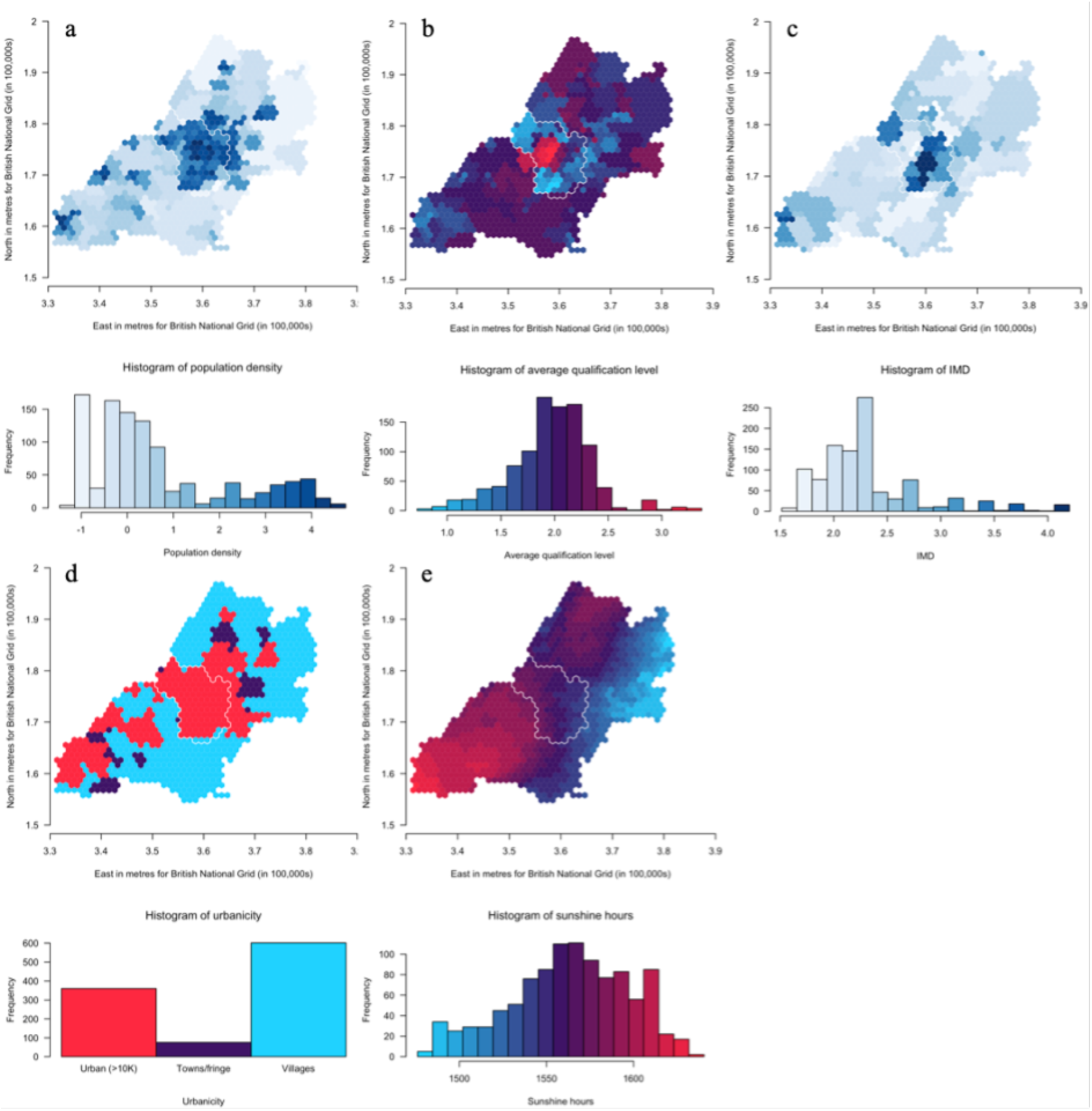
Maps of population density (a), average qualification level (b), Index of Multiple Deprivation (IMD) (c), level of urbanicity (d) and hours of sunshine (e) (30-year annual average from 1981 to 2010) for the ALSPAC catchment area, in and around Bristol. *The maps in the figures show a) log transformed population density (from 2001 census data) ranging from low (light blue) to high (dark blue), b) average qualification level (from 2001 census data) ranging from low (blue) to high (red), c) log transformed Index of Multiple Deprivation (IMD) (from 2000) ranging from low (light blue) to high (dark blue), d) level of urbanicity (from 2001 census data) showing urban i.e., with a population greater than 10,000 (red), towns/fringe areas which included an settlement area classified as part of a small town or urban fringe (purple) and villages, which included dispersed dwellings, hamlets and villages (blue), the latter two classifications determined based on household densities and e) hours of sunshine (30-year annual average from 1981 to 2010) ranging from low (blue) to high (red). Histograms shows the distribution of the respective measures coloured in the same way*.

### Polygenic scores and participation and migration measures

There was strong evidence for a negative association of child’s ADHD PGS with child’s participation (ß=-0.30; 95% CI=-0.45, -0.15; *p*=9.05×10^-05^) and mother’s ADHD PGS with mother’s participation (ß=-0.35; 95% CI=-0.54, -0.16; *p*=2.72×10^-04^) (Supplementary Table S7). We did not find strong evidence of associations of either autism or ADHD PGS with the migration measures (see Supplementary Table S8).

## Discussion

We found spatial variation in genetic influences for both autistic and ADHD traits measured using PGS in a single city region. This corroborates previous research using twin analysis that identified spatial variation in the genetic influence of autistic traits on a national scale (Reed et al., 2021). Our results were consistent across different *pT* for social autistic traits, but less consistent for the autistic traits mean factor score and ADHD traits.

This spatial variation in genetic influence on autism and ADHD traits (by which we mean the association between autism or ADHD PGS and these traits) supports interplay between genetic influence and geographical environments, indicative of gene-environment interactions or correlations. Despite the expected low predictive power of the PGS, the association between PGS for autism and ADHD with their phenotypic counterparts does vary spatially. This suggests that certain geographically distributed environments draw out or mask genetic influences on autism and ADHD. This highlights the importance of local context when conducting PGS studies, going beyond the typical population-level analyses. However, it is difficult to identify consistent patterns across the *pT* for ADHD. We note that confidence intervals for each point overlap with those of the corresponding points on the other *pT* maps, so despite appearances the maps are not necessarily inconsistent. This consistency may become clearer as GWAS of larger samples identify variants associated with autism and ADHD that explain a greater amount of variance in the phenotypes.

We investigated specific environmental characteristics that may be correlated with this spatial variation in associations between PGS and respective traits. We found strong evidence of spatial correlation between the variation in these PGS associations and environmental characteristics that had previously been associated with population prevalence, with the exception of population density for the autistic traits mean factor score, and urbanicity for social autistic traits. Many of these environmental characteristics are correlated, so the consistency of associations is reassuring. The relationships we observe with qualification level and IMD suggest that area level education and SEP may amplify genetic influences on these neurodevelopmental traits. This fits with previous phenotypic literature suggesting their prevalence is correlated with SEP (Bakian et al., 2015; Hoffman et al., 2012; Russell et al., 2015). Qualification level tends to be higher in less deprived areas, so the fact we observe opposite correlations for these with the PGS association maps fits with this relationship.

We observe a strong relationship between the maps for PGS associations with social autistic traits and hours of sunshine, where there is greater genetic influence in areas with more sunshine. This may be linked to previous reports of a relationship between decreased vitamin D levels and increased prevalence of autism (Cannell, 2017; Hastie et al., 2019; B. K. Lee et al., 2019; Vinkhuyzen et al., 2018). Despite a similar association for prevalence (Arns et al., 2013), the correlation between maps for PGS associations with ADHD traits and annual sunshine was in the opposite direction.

This is not inconsistent, because influences on prevalence and aetiology are not necessarily the same. But if true it would suggest a different mechanism of action for autistic and ADHD traits. However, as noted earlier, the maps for ADHD are not strongly correlated across thresholds (unlike for social autistic traits), so the results for ADHD should be interpreted with caution. We also found negative correlations between maps of genetic influence and maps of population density and urbanicity, suggesting that the genetic variants are more predictive of these traits in rural areas. Alongside previous findings of higher prevalence for autism in more urban and densely populated areas (Marlene B. Lauritsen et al., 2014; Marlene Briciet Lauritsen et al., 2005; Wu & Jackson, 2017), this might suggest that the impact of urban living is a more direct environmental effect that makes genetic variation relatively unimportant.

### Limitations

Whilst we observe spatial variation in genetic influences in this study, there are a few points to consider when interpreting these results. We found that greater polygenic risk for ADHD was associated with decreased participation for both children and mothers, in line with a previous study in ALSPAC (A. E. Taylor et al., 2018).

Therefore, analyses including the ADHD PGS may be biased by selection on study participation, which could result in distorted estimates. The child participation measures will partially capture mother’s participation as well (e.g., child-based questionnaires completed by mothers), but due to the age we examined, there were few measures available that were completed by the child. Similarly, migration could plausibly occur due to underlying genetic risk for a trait in parents, which in turn could influence the spatial patterning for offspring genetic risk. However, as we do not find evidence of this, it is unlikely that this will be having a large effect on our findings. The ADHD PGS explained more variance in ADHD traits than the autism PGS with social autistic traits. This is in line with the phenotypic variance explained in the original articles: 5.5% for ADHD compared to 2.5% for autism. However, the variance explained in our study was very low, so the spatial variation in genotype may not reflect spatial variation in the phenotype for this reason. This is likely to improve as GWAS become larger and more powered and future studies should explore this further.

Although both autistic traits and ADHD vary in presentation across the lifespan, most of our measures were obtained at age 10. However, autism and ADHD are neurodevelopmental conditions with traits arising early so it is likely these will be apparent by the time of measurement. The autistic traits mean factor score also addresses this issue by incorporating measures taken from a range of time points throughout childhood. However, this mean factor score also has its own limitations. For example, because it is an average score it assumes that the measures all explain equal amounts at each time point, so we are assuming a lifetime spatial measurement invariance, which may not be the case. Additionally, the correlation with the SCDC measure is not strong, likely due to the measures capturing different aspects of autistic traits.

There will be measurement error in the estimated effect sizes for individual genetic variants used to construct the PGS, which may reduce precision (Dudbridge, 2013), although the discovery GWAS samples were large, which helps to mitigate this issue. Similarly, there is likely to be a mixture of true and false positive associations in GWAS with many genome-wide significant hits. Modelling associations for a range of *pT* and observing consistent patterns, as we have, helps overcome this potential issue (Maher, 2015). Our analyses were conducted in a population sample of European ancestry, so results may not generalise to populations from other ancestral backgrounds (De La Vega & Bustamante, 2018). Assortative mating is also thought to be more common in autism (Nordsletten et al., 2016), which may bias autism GWAS and therefore make the PGS less accurate (Brumpton et al., 2020). However, the exact impact this would have on our results is not clear.

## Conclusion

In summary, our results demonstrate spatial variation in known genetic influences for both autism and ADHD traits in a single city region. This variation is associated with some of the environmental factors that are also associated with prevalence. Future research might examine these associations further along with a wider range of environmental variables. We hope that mapping the landscape of genetic influences may aid the identification of new spatially distributed environments that moderate genetic influences on autistic traits or ADHD. Identifying these factors and how they interact could one day lead to social policy interventions to improve outcomes for those with these developmental traits.

## Data Availability

Data availability
ALSPAC data access is through a system of managed open access. Access can be applied for as detailed in the ALSPAC access policy.
Code availability
The analysis code used in this study is available upon request from the authors.

## Acknowledgements

We are extremely grateful to all the families who took part in this study, the midwives for their help in recruiting them, and the whole ALSPAC team, which includes interviewers, computer and laboratory technicians, clerical workers, research scientists, volunteers, managers, receptionists and nurses.

