## Supporting Information for "Mapping associations of polygenic scores with autism and ADHD traits in a single city region"

**Supplementary methods**

*1. Autistic traits mean factor score*

The autistic traits mean factor score was derived from 93 measures originally taken across a number of time points from 6 months old up to age 9, and including, amongst other measures, the Social and Communication Disorders Checklist (SCDC). Missing data were imputed using data from the other 92 measures where necessary and then principal factor analysis was conducted, details of which can be found in the original article ^1^. The mean factor score was calculated from the top 7 factors from the analysis and was found to have the strongest association with autism in the previous study and therefore we have chosen to use it here. The factor mean score ranges from -3.62 to 0.70 in this study and a more negative score corresponds with a higher measure of autistic traits. Therefore, for ease of interpretation, we flipped the sign of the resulting effect estimates by multiplying them by -1. The distribution of the raw factor scores for this measure can be seen in supplementary fig 3.

*2. Location data*

This covers the city of Bristol (population N=463,000), North Somerset (including Weston Super Mare, population N=88,000), South Gloucestershire and a section of Bath & North East Somerset (but excluding the city of Bath). In 2001 (the timepoint of interest in this study) this area had a total population of approximately one million ^2^. These target locations were centroids of hexagons in a grid-like formation across a map of this region (Supplementary fig 4). There were a total of 1,036 target locations.

To preserve participants’ anonymity, locations were matched, and distances calculated by the ALSPAC team, who returned an anonymised dataset for each location to the researchers for analysis.

*3. Genetic data*

Samples for children were genotyped using the Illumina HumanHap 550 quad chip. Genome-wide data for children were generated by Sample Logistics and Genotyping Facilities at the Wellcome Trust Sanger Institute and LabCorp (Laboratory Corporation of America) using support from 23andMe. Quality control measures were used, and individuals were excluded based on gender mismatches, minimal or excessive heterozygosity, disproportionate levels of individual missingness (>3%) and insufficient sample replication (identity by descent [IBD] <0.8). Population stratification was assessed by multidimensional scaling analysis and compared with Hapmap II (release 22) European descent (CEU), Han Chinese, Japanese and Yoruba reference populations; all individuals with non-European ancestry were removed. SNPs with a minor allele frequency (MAF) of <1%, a call rate of <95% or evidence of violations of Hardy-Weinberg equilibrium (P<5x10^-07^) were removed. Cryptic relatedness was measured as the proportion of IBD>0.1.

Genotyping for mothers was conducted ysing the Illumina human660W-quad array at Centre National de Génotypage (CNG) and genotypes were called with Illumina GenomeStudio. Quality control measures were used. SNPs were removed if they displayed more than 5% missingness and a Hardy-Weinberg equilibrium P value of <1x10^-06^. SNPs with a MAF <1% were removed. Samples were excluded if they displayed >5% missingness, had indeterminable X chromosome heterozygosity or extreme autosomal heterozygosity. Samples showing evidence of population stratification were identified by multidimensional scaling of genome-wide identity by state pairwise distances using the four HapMap populations as reference, and then excluded. Cryptic relatedness was assessed using an IBD estimate >0.125, which is expected to correspond to approximately 12.5% alleles shared IBD or a relatedness at the first cousin level.

Related individuals that passed all other quality control thresholds were retained during subsequent phasing and imputation. 9,115 children and 500,527 SNPs passed these. 9.048 mothers and 526,688 SNPs passed these filters. 477,482 SNP genotypes in common between mothers and children were removed. SNPs with genotype missingness >1% were also removed. 321 individuals were removed due to potential ID mismatches. Haplotypes were estimated using ShapeIT (V2.r644) which utilises relatedness during phasing. A phased version of the 1000 genomes reference panel (Phase 1, version 3) was obtained from the Impute2 reference data repository (phased using ShapeIT v2.r644, haplotype release date Dec 2013). Imputation of the target data was performed using Impute V2.2.2 against the reference panel using all 2186 reference haplotypes (including non-Europeans).

This resulted in 8,237 eligible children and 8.196 eligible mothers with available genotype data.

*4. Polygenic score construction*

To create polygenic scores for autism and ADHD we used genetic data in ALSPAC filtered by an imputation score of 0.8 and a minor allele frequency of 0.01. After this filtering and selecting SNPs where genotype data was available in ALSPAC, SNPs were clumped for linkage disequilibrium, using the –clump command in Plink and an R^2^ of 0.1. We generated weighted polygenic scores for each phenotype by summing the number of risk alleles present for each SNP (0, 1 or 2) weighted by the beta of that SNP from the discovery samples for ADHD ^3^ and autism ^4^, using the –score command in Plink. We present results for polygenic scores constructed at p < 0.5, 1x10^-5^, 5x10^-8^ thresholds in the discovery GWAS and for the threshold that explained the most variance for each of the phenotypes in ALSPAC. We did this so that we could see whether results were consistent across p value thresholds. We chose thresholds that included many SNPs with the 0.5 pT, genome-wide significant SNPs in the 5x10^-8^ pT and a more lenient threshold than genome-wide with the 1x10^-5^ pT. Polygenic scores were z-standardised; therefore, results can be interpreted as per standard deviation (SD) increase in score. We additionally created polygenic scores for the mothers in ALSPAC for autism and ADHD using the same p-value thresholds.

*5. Unweighted polygenic score analyses*

We conducted analyses which were unweighted to obtain estimates for the association of unweighted polygenic scores with the phenotype. This allows us to compare estimates between unweighted and weighted analyses. All statistical analyses were conducted in R (version 3.5.1) (R Team 2016). We used linear regression models, adjusted for age, sex and 20PCs of population structure, to assess these associations. We used the R^2^ to find the pT that explained the most variation in each phenotype, out of 13 different polygenic scores and used these for subsequent analyses as described above, along with the other pT polygenic scores.

*6. Environmental measures*

Population density (as density per hectare) and average qualification level and the level of urbanicity (urban >10,000 population, towns/fringe areas and villages or isolated dwellings) from 2001 census data for Lower Super Output Areas (LSOAs) ^6^, the index of multiple deprivation (IMD) from the year 2000 (where a higher number indicates a more deprived area, available for download from the National Archives) and hours of bright sunshine as a 30 year annual average (from 1981 to 2010, available for download here: <https://www.metoffice.gov.uk/research/collaboration/ukcp/download-data>). Data were assigned to the same hexagonal grids that were used for the polygenic score maps to allow for comparisons. Values for these variables were assigned based on where the centroid of each hexagon fell within the different output areas.

*7. Participation and migration measures*

We created separate measures of each child’s and each mother’s participation in ALSPAC, up to child age 11. For children, this was the number of child-focussed and child-completed questionnaires returned plus the number of clinic visits attended (total possible score of 36). For mothers, this was the number of child-focussed questionnaires completed plus the number of questionnaires that the mother responded to about herself (total possible score 34). To measure migration, we had data available on whether a family had moved within Avon or outside of Avon, up to age 10. We constructed a binary variable for this to indicate whether they had moved out of Avon. The same variable was used for children and mothers.

**Supplementary figures**

Supplementary Fig S1. Distribution of ADHD traits as measured using the SDQ questionnaire in the ALSPAC cohort

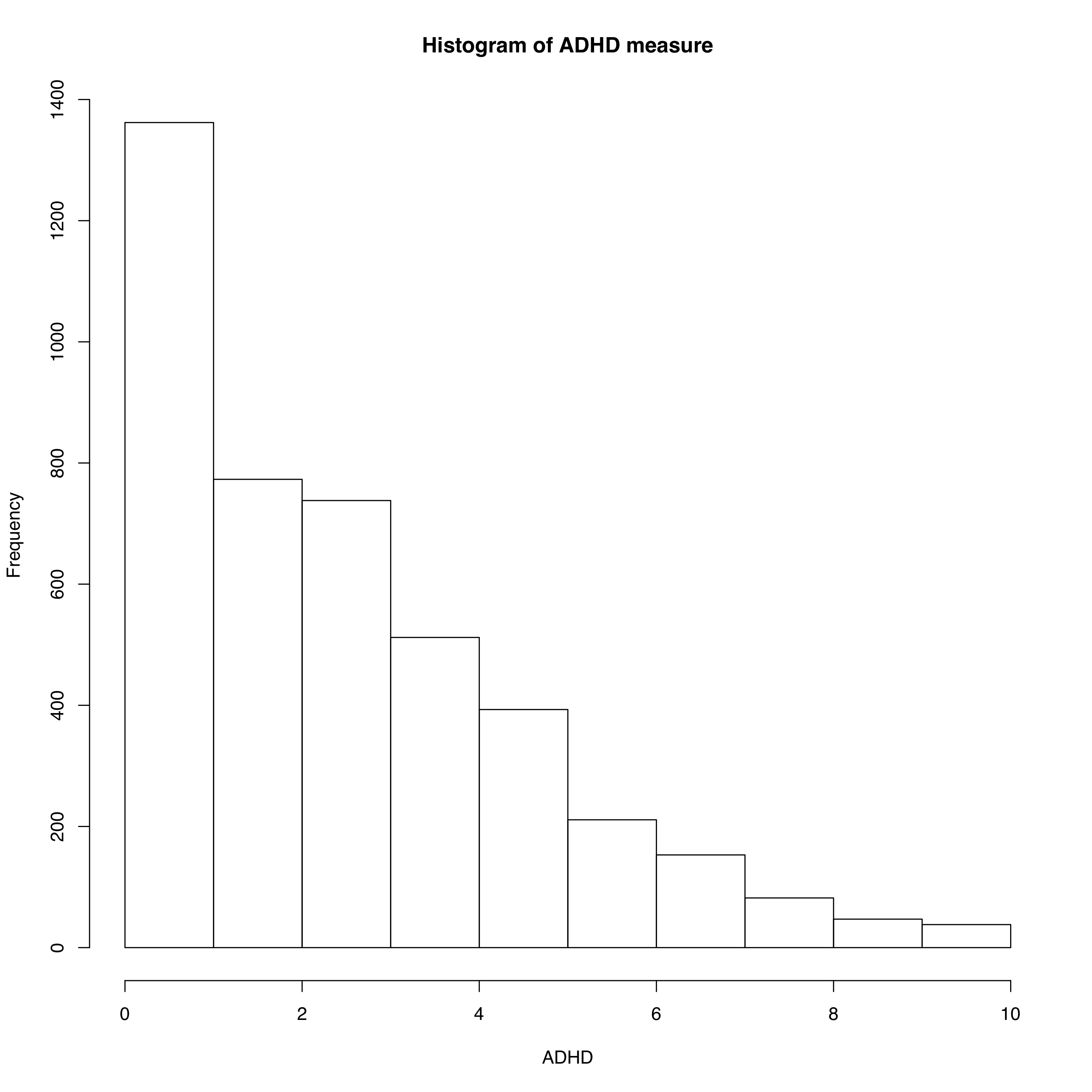

Supplementary Fig S2. Distribution of social autistic traits as measured using the SCDC questionnaire in the ALSPAC cohort

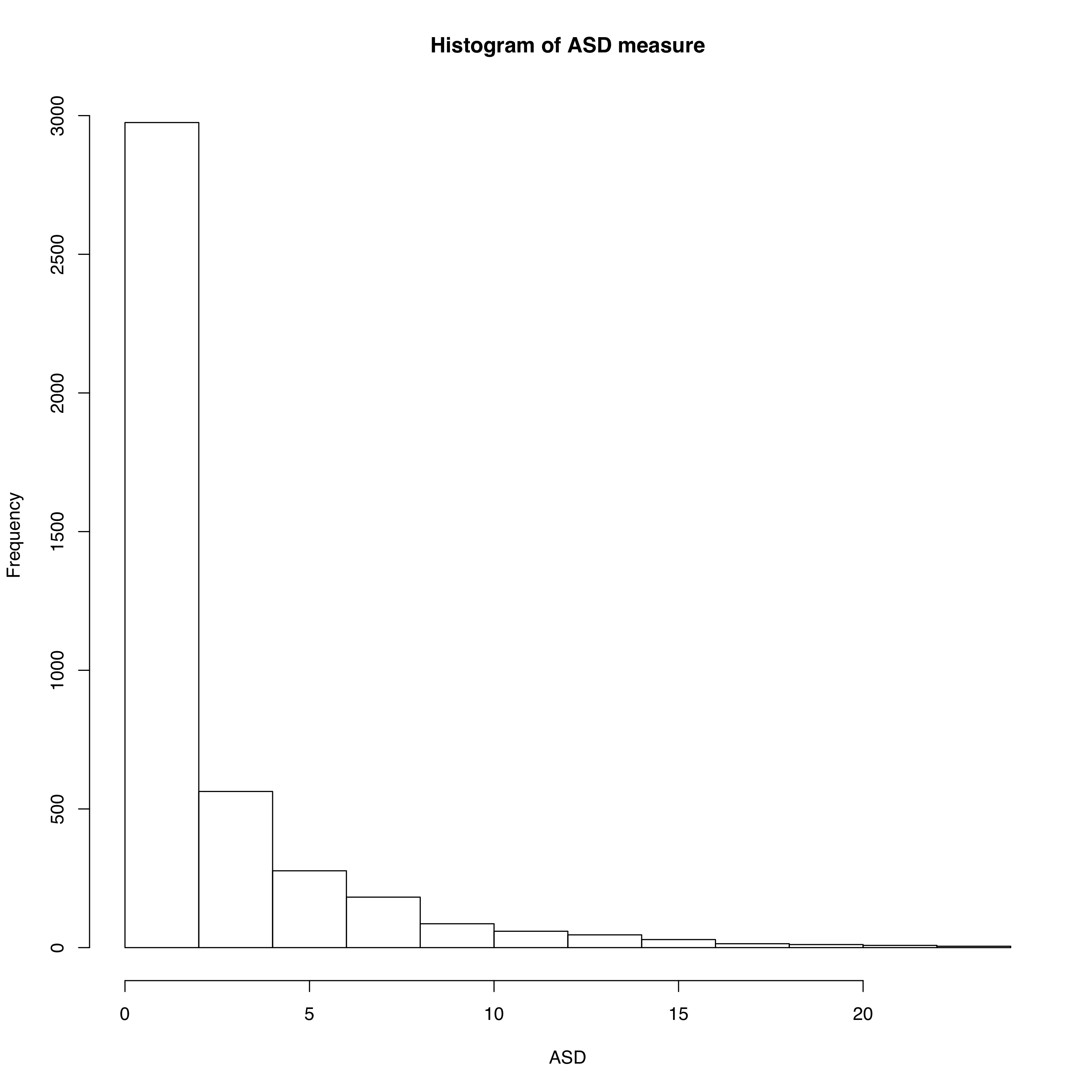

Supplementary Fig S3. Distribution of the autistic traits mean factor score in the ALSPAC cohort

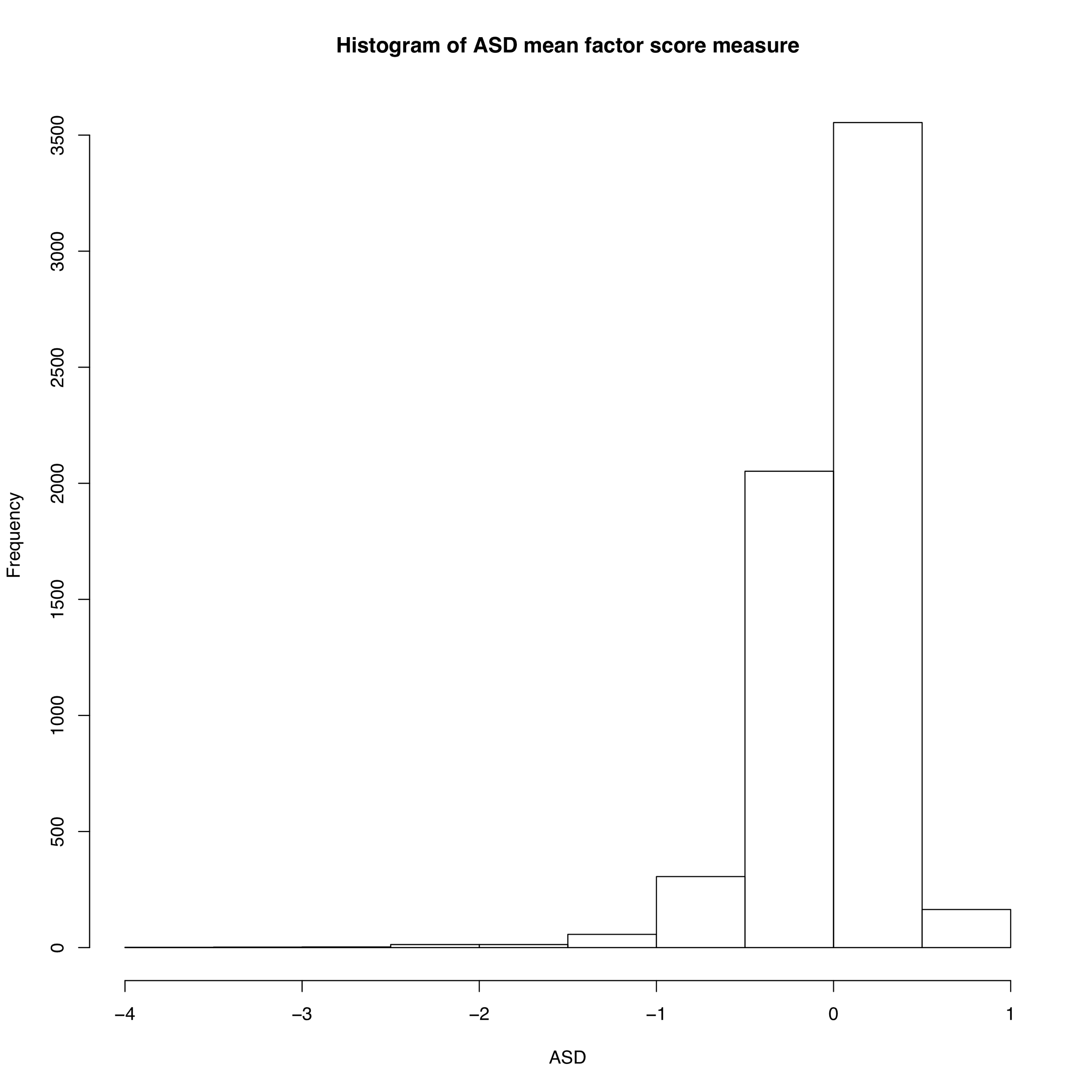

Supplementary Fig S4. Hexagonal grid of target locations across for analysis across Bristol and surrounding areas, based on three health districts (Southmead, Frenchay and Bristol and Weston).

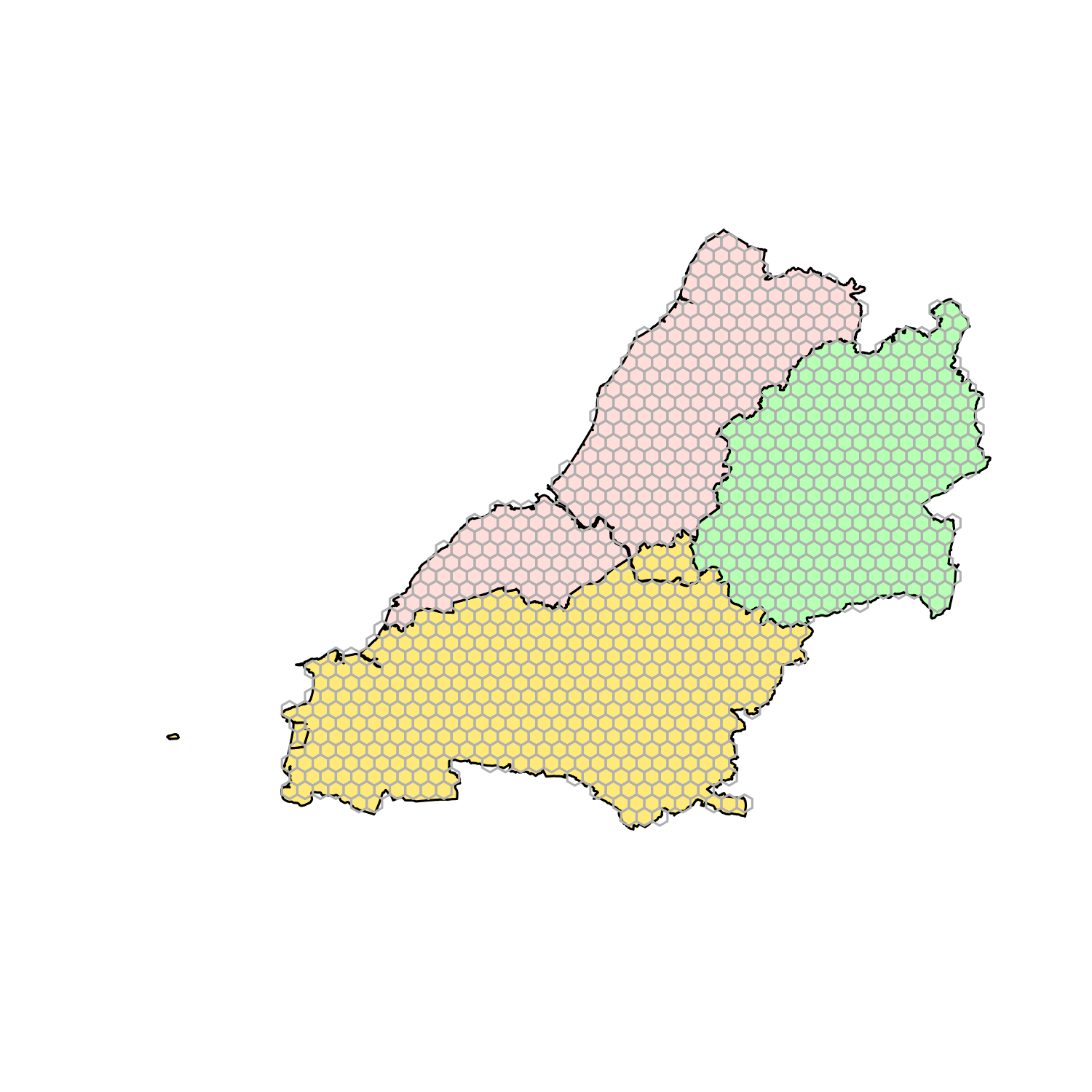

**Frenchay**

**Southmead**

**Bristol and Weston**

Supplementary Fig S5. Maps showing the association of the polygenic score for ADHD (using the European GWAS results) with ADHD traits

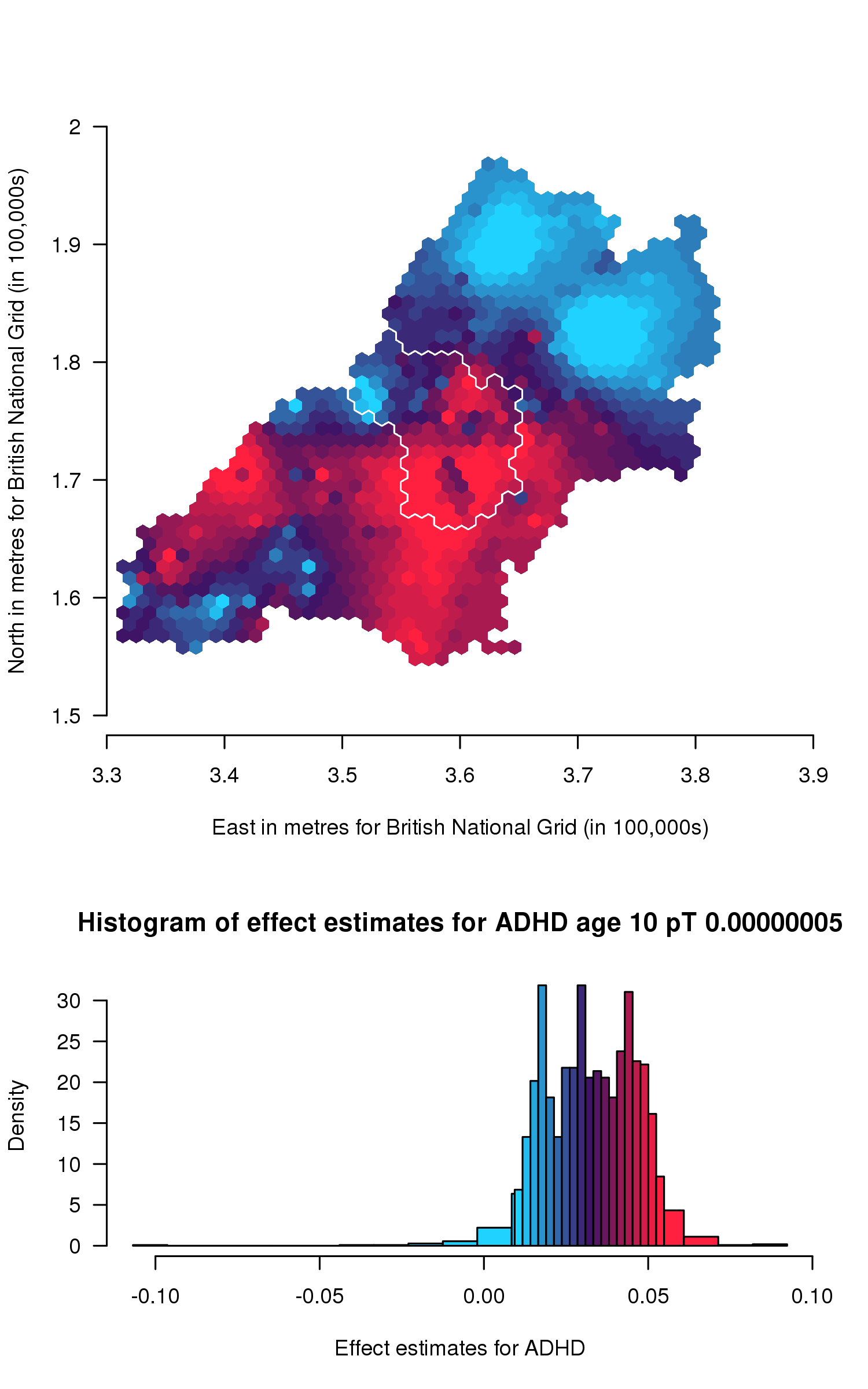

a

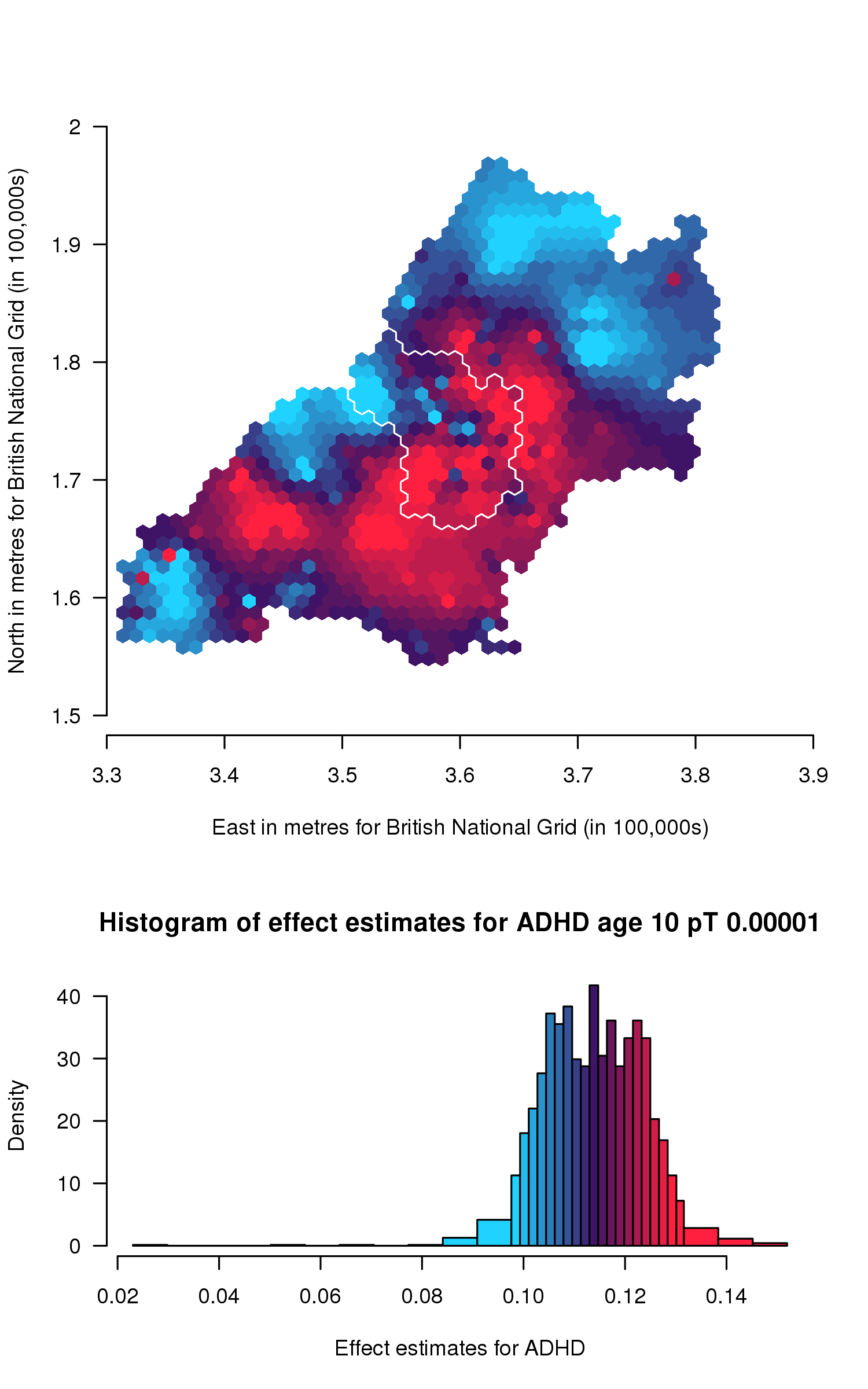

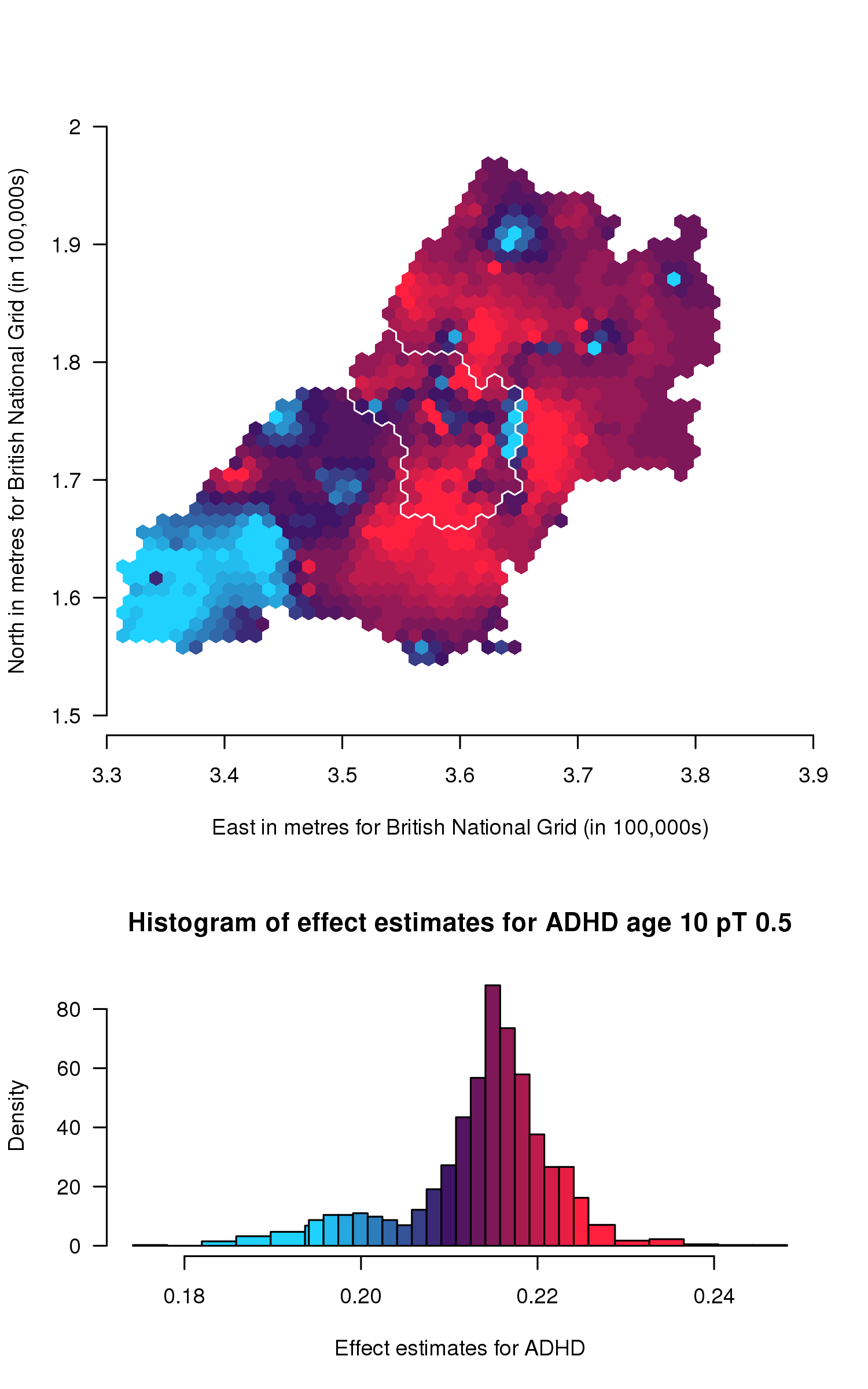

b

c

*Spatial variation in genetic influences ranging from low (blue) to high (red). Histograms show the distribution of effect estimates, coloured in the same way.*

**Supplementary tables**

Supplementary Table S1. Descriptive statistics for participants included in analyses for social autistic traits, autism mean factor score and ADHD traits.

| **Trait measure** | **Median trait score (IQR)** | **Mean age at measurement (SD)** | **N** | **Percentage male** |
| --- | --- | --- | --- | --- |
| Social autistic traits | 1.00 (0.00-3.00) | 10.72 (0.12) | 4,255 | 49.87% |
| Autistic traits mean factor score | 0.08 (-0.14-0.25) | Cross-age composite | 6,165 | 50.84% |
| ADHD traits | 3.00 (1.00-4.00) | 9.64 (0.12) | 4,309 | 50.24% |

*ADHD=Attention Deficit Hyperactivity Disorder; IQR = Inter Quartile Range; SD = Standard Deviation.*

Supplementary Table S2. Polygenic score analyses for ADHD traits for each of the 13 polygenic scores (N=5,258).

|  | **Adjusting for age and sex** | | | **Adjusting for age, sex and 20 PCs** | |
| --- | --- | --- | --- | --- | --- |
| **Polygenic score threshold** | **Beta (95% CI)** | **P-value** | **R^2^*** | **Beta (95% CI)** | **P-value** |
| 0.00000005 | 0.06 (-0.002, 0.12) | 0.06 | 0.00080 | -0.03 (-0.09, 0.03) | 0.29 |
| 0.0000001 | 0.04 (-0.01, 0.10) | 0.14 | 0.00058 | -0.02 (-0.08, 0.04) | 0.54 |
| 0.000001 | 0.11 (0.05, 0.17) | 3.99x10^-04^ | 0.0026 | -0.01 (-0.07, 0.04) | 0.64 |
| 0.00001 | 0.09 (0.03, 0.15) | 1.89x10^-03^ | 0.0019 | -0.01 (-0.07, 0.05) | 0.85 |
| 0.0001 | 0.10 (0.04, 0.16) | 7.74x10^-04^ | 0.0019 | 0.01 (-0.05, 0.07) | 0.66 |
| 0.001 | 0.16 (0.10, 0.22) | 7.12x10^-08^ | 0.0052 | 0.03 (-0.03, 0.09) | 0.29 |
| 0.01 | 0.20 (0.14, 0.26) | 7.40x10^-11^ | 0.0075 | 0.06 (-0.001, 0.12) | 0.06 |
| 0.05 | 0.23 (0.17, 0.29) | 5.30x10^-14^ | 0.010 | 0.01 (-0.05, 0.07) | 0.01 |
| 0.1 | 0.23 (0.17, 0.29) | 4.14x10^-14^ | 0.010 | 0.02 (-0.02, 0.10) | 0.02 |
| 0.2 | 0.23 (0.17, 0.29) | 7.48x10^-14^ | 0.0099 | 0.04 (-0.02, 0.10) | 0.04 |
| 0.3 | 0.23 (0.17, 0.29) | 2.22x10^-14^ | 0.010 | 0.04 (-0.02, 0.10) | 0.04 |
| 0.4 | 0.23 (0.17, 0.29) | 1.37x10^-14^ | 0.011 | 0.04 (-0.02, 0.10) | 0.04 |
| 0.5 | 0.23 (0.18, 0.29) | 8.25x10^-15^ | 0.011 | 0.03 (-0.03, 0.09) | 0.03 |

*The effect estimate (beta) is from linear regression models between the polygenic score for ADHD and ADHD traits, adjusting for the covariates and represents the change in ADHD traits per SD increase in the polygenic score.*

^*^ *r-squared value from linear model with just the polygenic score as the exposure and no covariates included*

Supplementary Table S3. Polygenic score analyses for social autistic traits (from SCDC) for each of the 13 polygenic scores (N=5,200).

|  | **Adjusting for age and sex** | | | **Adjusting for age, sex and 20 PCs** | |
| --- | --- | --- | --- | --- | --- |
| **Polygenic score threshold** | **Beta (95% CI)** | **P-value** | **R^2^*** | **Beta (95% CI)** | **P-value** |
| 0.00000005 | 0.05 (-0.04, 0.15) | 0.27 | 0.00028 | -0.07 (-0.16, 0.03) | 0.17 |
| 0.0000001 | 0.05 (-0.04, 0.15) | 0.27 | 0.00028 | -0.07 (-0.16, 0.03) | 0.17 |
| 0.000001 | 0.02 (-0.08, 0.11) | 0.73 | 0.000038 | 0.006 (-0.09, 0.10) | 0.90 |
| 0.00001 | 0.01 (-0.08, 0.11) | 0.76 | 0.0000097 | -0.004 (-0.10, 0.09) | 0.94 |
| 0.0001 | 0.05 (-0.05, 0.14) | 0.33 | 0.00018 | -0.009 (-0.10, 0.09) | 0.86 |
| 0.001 | 0.05 (-0.05, 0.14) | 0.32 | 0.00018 | 0.04 (-0.05, 0.14) | 0.36 |
| 0.01 | 0.03 (-0.06, 0.12) | 0.52 | 0.000052 | 0.07 (-0.02, 0.16) | 0.14 |
| 0.05 | 0.05 (-0.05, 0.14) | 0.34 | 0.00017 | 0.04 (-0.06, 0.13) | 0.43 |
| 0.1 | 0.09 (-0.002, 0.19) | 0.06 | 0.00068 | 0.0001 (-0.09, 0.09) | 1.00 |
| 0.2 | 0.08 (-0.02, 0.17) | 0.11 | 0.00048 | -0.03 (-0.12, 0.07) | 0.56 |
| 0.3 | 0.08 (-0.01, 0.18) | 0.09 | 0.00051 | -0.03 (-0.12, 0.07) | 0.51 |
| 0.4 | 0.09 (-0.01, 0.18) | 0.07 | 0.00060 | -0.03 (-0.12, 0.06) | 0.54 |
| 0.5 | 0.09 (-0.004, 0.18) | 0.06 | 0.00065 | -0.03 (-0.12, 0.07) | 0.59 |

*The effect estimate (beta) is from linear regression models between the polygenic score for autism and social autistic traits, adjusting for the covariates and represents the change in social autistic traits per SD increase in the polygenic score.*

^*^ *r-squared value from linear model with just the polygenic score as the exposure and no covariates included*

Supplementary Table S4. Polygenic score analyses for the autistic traits mean factor score for each of the 13 polygenic scores (N=7,505).

|  | **Adjusting for age and sex** | | | **Adjusting for age, sex and 20 PCs** | |
| --- | --- | --- | --- | --- | --- |
| **Polygenic score threshold** | **Beta (95% CI)** | **P-value** | **R^2^*** | **Beta (95% CI)** | **P-value** |
| 0.00000005 | 0.004 (-0.004, 0.011) | 0.37 | 0.00016 | -0.008 (0.0007, 0.02) | 0.03 |
| 0.0000001 | 0.004 (-0.004, 0.011) | 0.37 | 0.00016 | -0.008 (0.0007, 0.02) | 0.03 |
| 0.000001 | -0.001 (-0.009, 0.006) | 0.72 | 0.0000024 | -0.003 (-0.005, 0.01) | 0.45 |
| 0.00001 | 0.001 (-0.007, 0.008) | 0.90 | 0.0000025 | -0.001 (-0.007, 0.009) | 0.81 |
| 0.0001 | 0.0001 (-0.008, 0.008) | 0.98 | 0.0000000097 | 0.003 (-0.01, 0.005) | 0.42 |
| 0.001 | 0.003 (-0.004, 0.01) | 0.38 | 0.00015 | -0.002 (-0.006, 0.009) | 0.67 |
| 0.01 | 0.008 (0.001, 0.02) | 0.03 | 0.00048 | -0.004 (-0.003, 0.01) | 0.26 |
| 0.05 | 0.01 (0.002, 0.02) | 0.02 | 0.00076 | -0.003 (-0.005, 0.01) | 0.49 |
| 0.1 | 0.01 (0.002, 0.02) | 0.01 | 0.00091 | -0.002 (-0.005, 0.01) | 0.55 |
| 0.2 | 0.01 (0.004, 0.02) | 0.004 | 0.0011 | -0.001 (-0.007, 0.009) | 0.85 |
| 0.3 | 0.01 (0.003, 0.02) | 0.006 | 0.00098 | -0.003 (-0.005, 0.01) | 0.50 |
| 0.4 | 0.01 (0.003, 0.02) | 0.006 | 0.00099 | -0.002 (-0.006, 0.009) | 0.68 |
| 0.5 | 0.01 (0.004, 0.02) | 0.003 | 0.0012 | -0.002 (-0.006, 0.01) | 0.66 |

*The effect estimate (beta) is from linear regression models between the polygenic score for autism and the autistic traits mean factor score, adjusting for the covariates and represents the change the the autistic traits mean factor score per SD increase in the polygenic score.*

^*^ *r-squared value from linear model with just the polygenic score as the exposure and no covariates included*

Supplementary Table S5. Lee statistic test results for polygenic score maps across different p-value thresholds, using Monte-Carlo simulation with 10,000 permutations.

|  | **Monte-Carlo simulation of Lee’s L statistic, with 10,000 permutations (p-value)** |
| --- | --- |
| **ADHD traits (5x10^-8^ and 1x10^-5^)** | 0.14 (p=0.002) |
| **ADHD traits (1x10^-5^ and 0.5)** | -0.008 (p=0.07) |
| **Social autistic traits (5x10^-8^ and 1x10^-5^)** | 0.57 (p<2.00x10^-04^) |
| **Social autistic traits (1x10^-5^ and 0.1)** | 0.61 (p<2.00x10^-04^) |
| **Social autistic traits (0.1 and 0.5)** | 0.81 (p<2.00x10^-04^) |
| **Autistic traits mean factor score (5x10^-8^ and 1x10^-5^)** | 0.07 (p<2.00x10^-04^) |
| **Autistic traits mean factor score (1x10^-5^ and 0.5)** | -0.22 (p<2.00x10^-04^) |

*ADHD=Attention Deficit Hyperactivity Disorder*

Supplementary Table S6. Lee statistic test results for environmental risk factor maps compared to polygenic score maps for the score with a p-value threshold that explains the most variance.

|  | **Monte-Carlo simulation of Lee’s L statistic, with 10,000 permutations (p-value)** | | | | |
| --- | --- | --- | --- | --- | --- |
|  | **Log transformed population density** | **Average qualification level** | **Log transformed IMD** | **Urbanicity level** | **Sunshine hours (annual average)** |
| **ADHD traits (pT 0.5)** | -0.04 (p<2.00x10^-04^) | 0.07 (p<2.00x10^-04^) | -0.10 (p<2.00x10^-04^) | -0.14 (p<2.00x10^-04^) | -0.47 (p<2.00x10^-04^) |
| **Social autistic traits (pT 0.1)** | -0.11 (p<2.00x10^-04^) | 0.07 (p<2.00x10^-04^) | -0.18 (p<2.00x10^-04^) | -0.02 (p=0.02) | 0.57 (p<2.00x10^-04^) |
| **Autistic traits mean factor score (pT 0.5)** | 0.001 (p=0.91) | 0.13 (p<2.00x10^-04^) | -0.19 (p<2.00x10^-04^) | -0.05 (p<2.00x10^-04^) | -0.12 (p<2.00x10^-04^) |

*ADHD=Attention Deficit Hyperactivity Disorder, pT= p-value thresholds, IMD= indices of multiple deprivation*

Supplementary Table S7. Associations between mother and child polygenic scores for autism and ADHD and participation and migration measures.

|  | **Beta^1^ (95% CI)** | ***p*-value** |
| --- | --- | --- |
| *Participation* | | |
| Child’s participation ~ Child autism PGS | -0.11(-0.25, 0.04) | 0.17 |
| Child’s participation ~ Child ADHD PGS | -0.30 (-0.45, -0.15) | 9.05x10^-05^ |
| Mother’s participation ~ Mother autism PGS | -0.05 (-0.24, 0.14) | 0.61 |
| Mother’s participation ~ Mother ADHD PGS | -0.35 (-0.54, -0.16) | 2.72x10^-04^ |
| *Migration* | | |
| Migration out of Avon ~ Child autism PGS | 0.98 (0.91, 1.06) | 0.58 |
| Migration out of Avon ~ Child ADHD PGS | 0.94 (0.87, 1.01) | 0.09 |
| Migration out of Avon ~ Mother autism PGS | 1.00 (0.94, 1.07) | 0.92 |
| Migration out of Avon ~ Mother ADHD PGS | 0.94 (0.87, 1.00) | 0.06 |

*^1^Beta coefficient for linear regression models with participation measure or odds ratio for logistic regression models with migration measures.*

*ADHD=Attention Deficit Hyperactivity Disorder, PGS=polygenic score, CI=confidence interval*

Supplementary Table S8. Mean participation scores for children and mothers by migration group.

|  | **Mean participation score (SD)** |
| --- | --- |
| *Child participation score* | |
| Family did not move | 22.66 (12.84) |
| Family moved within Avon area | 23.12 (12.29) |
| Family moved outside Avon area | 23.85 (12.21) |
| *Mother participation score* | |
| Family did not move | 24.50 (10.95) |
| Family moved within Avon area | 24.81 (10.51) |
| Family moved outside Avon area | 25.95 (9.93) |

*SD=Standard deviation*

Supplementary Table S9. Summary of environmental measures used for ALSPAC participants

| **Measure** | **Output area** | **Range of data in the Bristol and surrounding areas or percentage (%) in each category** |
| --- | --- | --- |
| Population density (density per hectare) | Census lower super output area | 0.23 – 114.07 |
| Average qualification level | Census lower super output area | 0.72 – 3.37 |
| Level of urbanicity | Census lower super output area | Villages = 58.01%  Towns/fringe = 7.24%  Urban (>10k) = 34.75% |
| Indices of multiple deprivation | Ward | 4.55 – 66.80 |
| Hours of bright sunshine | 1-kilometre grid over United Kingdom | 1475.96 – 1640.55 |
